# Effects of oxycodone versus sufentanil on postoperative sleep quality and analgesia in patients after modified radical mastectomy: study protocol for a randomized, double-blind, controlled trial using wearable sleep monitoring

**DOI:** 10.64898/2026.05.20.26353683

**Authors:** Deng Qianyao, Hu Jie, Huang Lihua, Zheng Ju, Liang Zheng, Wu Ailing

## Abstract

**Background:** Postoperative sleep disorder, a frequently observed complication, is associated with heightened pain sensitivity, exacerbated inflammatory reactions, and compromised tissue repair. Sufentanil, a highly selective μ-opioid receptor agonist, is widely used in patient-controlled intravenous analgesia (PCIA) and has been associated with reduced sleep efficiency. Oxycodone, as a μ/κ dual receptor agonist, has shown a lower incidence of adverse effects in clinical practice. Despite these pharmacological differences, the comparative effects of oxycodone-versus sufentanil-based PCIA on postoperative sleep remain poorly characterized. Recent advances in wearable devices demonstrate strong agreement with polysomnography (PSG) in intergroup comparisons of sleep efficiency and total sleep time, enabling continuous, non-invasive, multi-night sleep monitoring and offering a viable alternative for clinical postoperative sleep research. Hence, we design this clinical trial to compare postoperative sleep efficiency between patients receiving oxycodone-based versus sufentanil-based PCIA under wearable sleep monitoring.

**Methods:** This study is a randomized, double-blind, placebo-controlled trial that was conducted at a single center. A sample size of 68 patients was determined through calculation, and these patients will be randomly assigned to either the oxycodone group or the sufentanil group. Sleep monitoring was initiated using a wristband device one day before surgery after recruitment. The sleep quality data at different setting time will be monitored. All patients will be followed up by blinded evaluators at baseline and 1, 2, and 30 days after the intervention. The follow-up included pain scores, postoperative complications and adverse events, etc.

**Discussion:** By integrating a modern photoelectric device with first-line analgesics, we hope the result of the study will inform perioperative sleep management, guide clinical analgesic selection, and improve patient recovery quality.

**Trial registration:** ID: ChiCTR2600118982, registered on February 13 2026

## Background

Perioperative sleep disorders (PSD) are an independent risk factor for postoperative delirium (POD), which is a common complication after surgery^1^. Radical mastectomy is a common surgical procedure in current clinical practice, with perimenopausal women aged 45-55 years in China representing the population with the highest incidence of this disease. Sufentanil and oxycodone are both commonly used first-line agents for postoperative analgesia. Studies have confirmed that traditional pure μ-receptor agonists, such as sufentanil, while providing analgesic effects, can significantly disrupt the normal sleep architecture^2^. This disruption is manifested as shortened deep sleep duration, decreased sleep efficiency, an increased number of awakenings, and impaired sleep continuity, thereby affecting the postoperative recovery process of patients.

Oxycodone, which acts on both μ and κ opioid receptors, may provide more balanced effects, and has been associated with improved postoperative recovery profiles and fewer adverse effects in randomized studies^3^. Study shows that dexmedetomidine combined with oxycodone can improve sleep quality^4^. Theoretically, this is more conducive to optimizing postoperative sleep quality. Despite these pharmacological differences, the comparative effects of oxycodone-versus sufentanil-based PCIA on postoperative sleep remain poorly characterized.

The normal sleep architecture is composed of non-rapid eye movement (NREM) sleep and rapid eye movement (REM) sleep. Among these, the N3 stage of NREM sleep, namely deep sleep (slow-wave sleep), plays a critical role in physical restoration, pain modulation, and immune function recovery^5^. The deep sleep period can significantly promote the secretion of growth hormone, reduce pain sensitivity, and improve inflammatory responses and tissue healing^6^. Concurrently, maintaining sleep continuity is a core element in ensuring sleep quality and reducing postoperative fatigue and adverse emotions^7^. Recent advances in wearable technology have enabled continuous, non-invasive monitoring of sleep-related parameters using integrated photoplethysmography (PPG) and actigraphy. At the group-comparison level, wearable-derived metrics such as sleep efficiency, total sleep time, nocturnal awakenings, heart rate variability, and respiratory rate demonstrate acceptable agreement with PSG for research applications^8^. These technologies provide an ecologically valid method to quantify postoperative sleep over multiple consecutive nights without interfering with clinical care.

Radical modified mastectomy involves soft tissue, with the surgical trauma being relatively limited to the operative field, resulting in relatively low analgesic requirements. The patients are generally physically stable with minimal interpatient variability and moderate postoperative pain burden, providing an appropriate clinical model to investigate opioid-related sleep effects. Given the current paucity of direct comparative studies on the effects of oxycodone versus sufentanil on postoperative sleep quality in breast cancer surgery, this study utilized a wearable device to objectively quantify sleep parameters. By comparing the effects of the two analgesic regimens on postoperative sleep duration, deep sleep, and sleep continuity, this research aims to provide evidence for optimizing multimodal analgesia protocols after breast cancer surgery and for refining postoperative analgesic strategies to enhance recovery.

## Methods

### Study design

This is a prospective, randomized, double-blind, parallel-group controlled clinical trial conducted at a single tertiary medical center. The study protocol has been approved by the Medical Ethical Committee of The Second People’s Hospital of Neijiang (MECSNJ202603) on January 30, 2026. This randomized double-blind, controlled clinical trial was registered at http://www.chictr.org.cn (ChiCTR2600118982) and will be conducted at the Department of Anesthesiology of The Second People’s Hospital of Neijiang. Participant recruitment and data collection are expected to be completed by November, 2026, results are expected by December 2026. As none of the above stages have been initiated, we hereby declare that all stages are still in an unstarted status in accordance with the journal’s requirements. Study duration per participant: from preoperative baseline night to postoperative day 3. The authors have completed the SPIRIT reporting checklist, Available at ***Figure 1. Figure 2*** gives the study flowchart that the patients will follow during the study and the flow diagram of participants is given in ***Figure 3***. This protocol is in accordance with the Standard Protocol Items: Recommendations for Interventional Trials (SPIRIT) 2013 statement^9,10^. We followed the SPIRIT checklist to address the recommended items in our clinical trial protocol and documents^9^.

**Figure 1.**
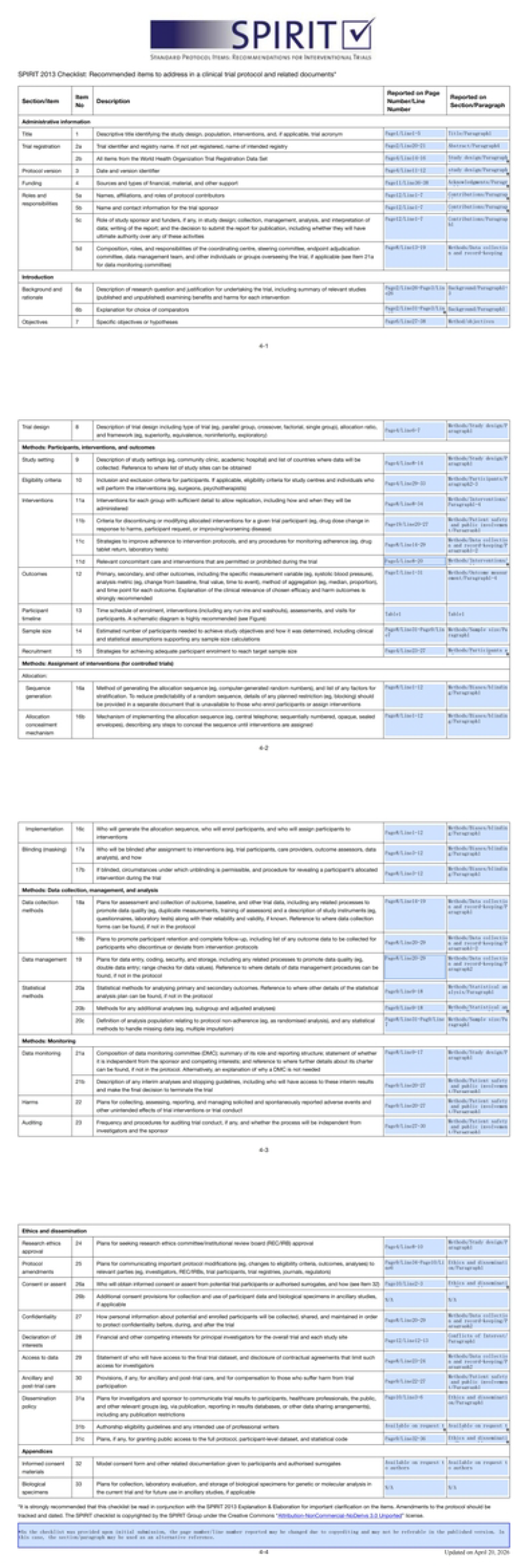
SPIRIT reporting checklist

**Figure 2.**
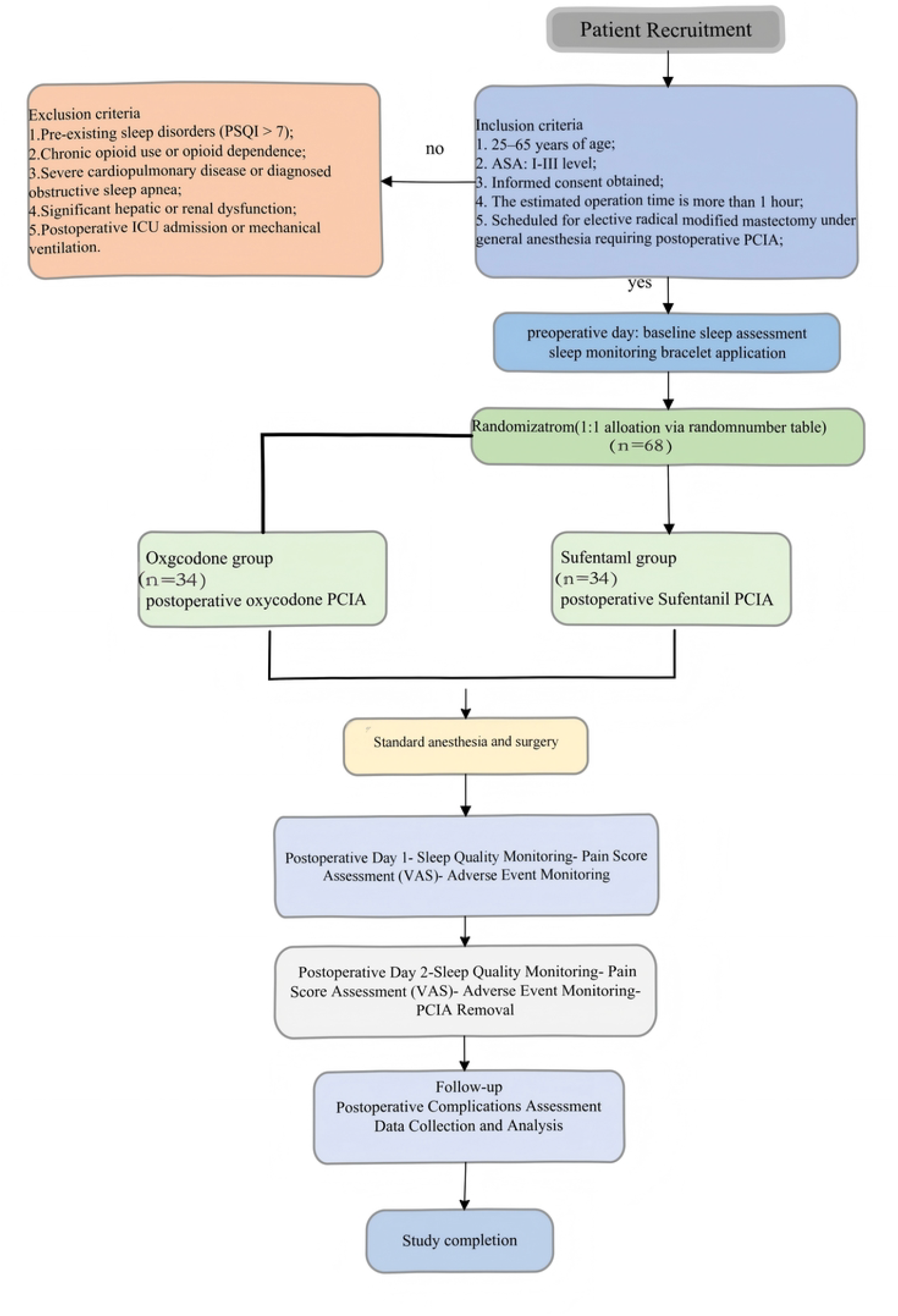
Flow diagram of participants

**Figure 3.**
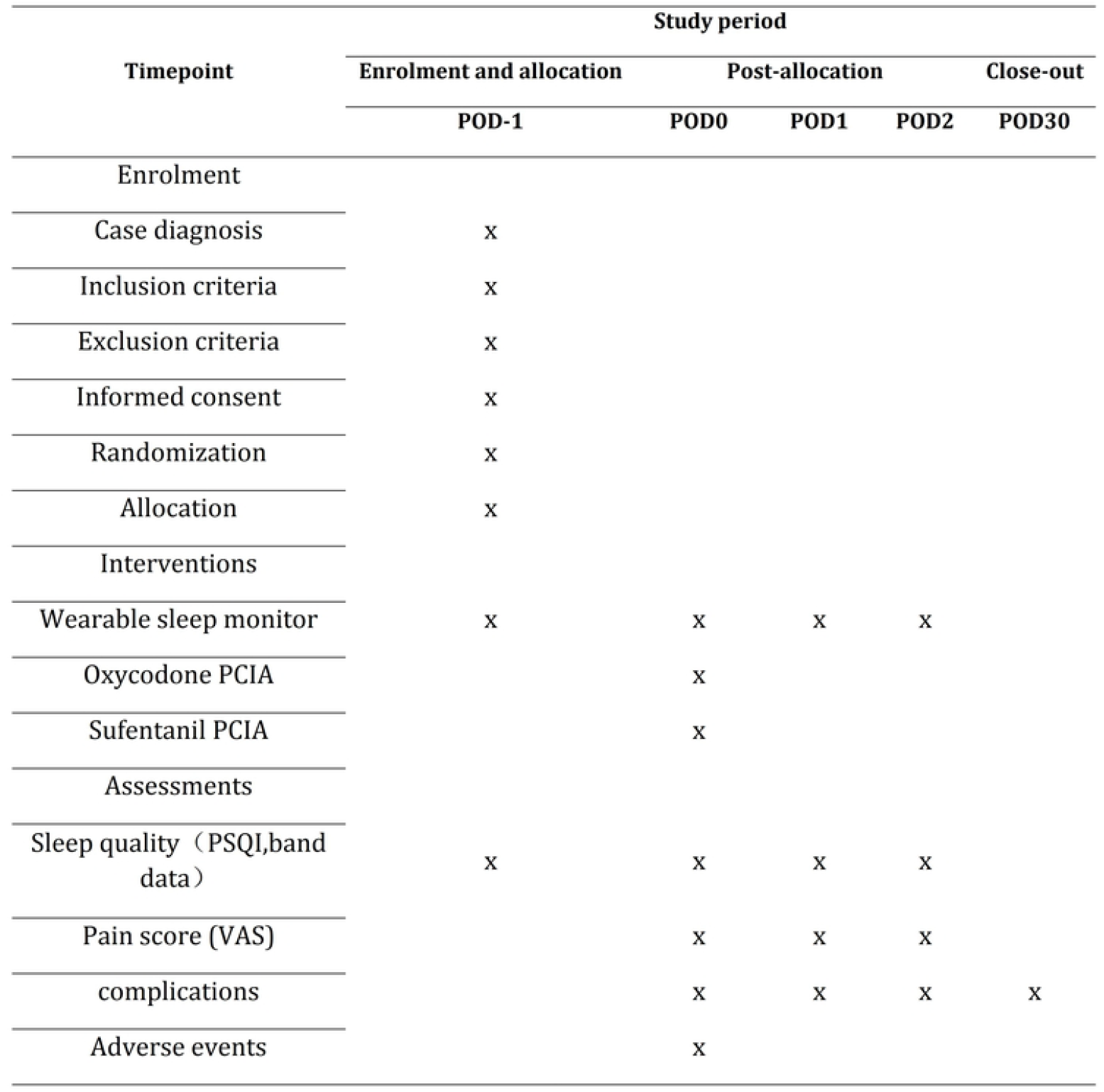
Schedule of recruitment, interventions, and assessments. According to SPIRIT 2013 statement: defining standard protocol items for clinical trials. Pod, postoperative day.

### Participants and recruitment

Through the screening of patients who have signed the informed consent form for elective surgery in The Second People’s Hospital of Neijiang, eligible patients will be recruited after signing the informed consent form for this study.

#### Inclusion criteria

1. 25–65 years of age;
2. ASA: I-III level;
3. Informed consent obtained;
4. The estimated operation time is more than 1 hour;
5. Scheduled for elective radical modified mastectomy under general anesthesia requiring postoperative PCIA;

#### Exclusion criteria

1. Pre-existing sleep disorders (PSQI > 7);
2. Chronic opioid use or opioid dependence;
3. Severe cardiopulmonary disease or diagnosed obstructive sleep apnea;
4. Significant hepatic or renal dysfunction;
5. Postoperative ICU admission or mechanical ventilation.

### Interventions

All participants will receive standardized general anesthesia. Anesthesia induction includes Sufentail 0.2-0.5ug/kg IV, propofol 1.5-2.5mg/kg IV, midazolam 1-2mg IV, and rocuronium 0.6-0.9mg/kg IV. Prophylactic use of dexamethasone and ondansetron. Intraoperative maintenance involves combined intravenous and inhalation anesthesia, with inhalation anesthesia using Sevoflurane at a concentration of 1% to 2%, and intravenous anesthesia including Propofol infusion at 1-2mg/kg/h for maintenance, and Remifentanil infusion at 0.1ug/kg/min for maintenance, with intraoperative monitoring of anesthesia depth to guide drug titration, strictly maintaining the BIS value between 40 and 60. Postoperatively, an Analgesia pump is connected, and non-steroidal anti-inflammatory drugs may be used if necessary.

- **Oxycodone group**: PCIA containing oxycodone at 0.6 mg/kg plus Ondansetron 8mg, diluted with normal saline to 100 ml, set the pump background speed to 4 ml/h, lock time 20 min, and the additional dose is 1 ml/h. Start with a loading dose of Oxycodone 0.05mg/kg via intravenous injection.
- **Sufentanil group**: PCIA containing sufentanil at 1.5 μg/kg plus Ondansetron 8mg, plus normal saline to 100 ml. set the pump background speed to 4 ml/h, lock time 20 min, and the additional dose is 1 ml/h. Start with a loading dose of sufentanil 0.1 μg/kg via intravenous injection.

All inhaled anesthesia will use the anesthetic machine produced by Ohmeda in the United States, and all intravenous anesthesia will use the medical microinjection pump compacts in Belan, Germany. All PCIA machine will use Aipeng medical analgesia infusion pump.

### Sleep Assessment Using Wearable Devices

Sleep was continuously monitored using a wrist-worn wearable device (Huawei Band 9, Huawei Technologies, China), which integrates photoplethysmography and tri-axial accelerometry. Sleep stages (wake, REM, light sleep, and deep sleep) were estimated using a validated proprietary algorithm based on heart rate variability (HRV) and actigraphy-derived features. Sleep efficiency, total sleep time, and nocturnal awakenings were extracted for analysis. Due to the bulky equipment and restricted patient mobility associated with conventional polysomnography (PSG), which may interfere with actual activity and rest, its routine clinical application remains limited. In this study, postoperative sleep was continuously monitored using a wrist-worn wearable device, which we conceptualize as a source of digital biomarkers reflecting real-world sleep physiology^11^. Digital biomarkers, defined as objective, quantifiable physiological and behavioral data collected through digital devices, offer a pragmatic approach to continuous monitoring in perioperative settings where conventional polysomnography (PSG) is not feasible. Importantly, the primary endpoint in this study—change in sleep efficiency—represents a composite measure of sleep continuity, integrating multiple aspects of sleep disruption (including sleep duration and wake after sleep onset). Compared with stage-specific metrics such as REM or deep sleep, sleep efficiency is less susceptible to misclassification errors inherent in wearable-based sleep staging algorithms, thereby enhancing robustness.

### Objectives

#### Primary Objective

To compare the change in postoperative sleep quality between patients receiving oxycodone-based versus sufentanil-based patient-controlled intravenous analgesia (PCIA).

#### Secondary Objectives

1. To compare postoperative pain intensity at rest and during movement.
2. To assess differences in sleep architecture parameters (total sleep time, nocturnal awakenings, REM and deep sleep proportions).
3. To evaluate autonomic and respiratory parameters derived from wearable devices.
4. To compare opioid consumption and the incidence of postoperative complications.

### Outcome Measures

#### Primary Outcome

1. Mean sleep efficiency across postoperative nights 1–2

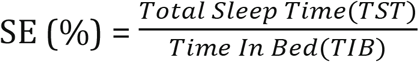
2. Mean sleep fragmentation index (SFI) across postoperative nights 1–2

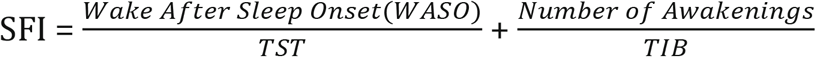

Based on the ward nurses’ lights-out schedule and the postoperative patients’ rest-activity patterns, TIB was defined as the interval from 22:00 to 7:00, and TST was defined as the total duration of sleep recorded by the wrist-worn device during the TIB period.

#### Primary Outcome Measures

1. Total Sleep Time (TST): actual sleep duration
2. Deep Sleep Time(N3): slow-wave sleep duration
3. Wake After Sleep Onset (WASO): total wakefulness time after sleep onset until final awakening.

#### Monitoring Periods

1. Preoperative baseline: 22:00 on the day before surgery to 07:00 on the day of surgery (drug-free, natural sleep).
2. Postoperative monitoring: 22:00 to 07:00 on postoperative day 1 (POD1) and postoperative day 2 (POD2).

#### Secondary Outcomes

1. Pain scores (VAS) at 6, 12, 24, and 48 hours postoperatively
2. Total opioid consumption within 48 hours
3. Wearable-derived heart rate, heart rate variability, and respiratory rate
4. Pittsburgh Sleep Quality Index (PSQI) scores and incidence of adverse events (respiratory depression, nausea/vomiting, excessive sedation, delirium)

### Biases/blinding

Randomization will be performed using a computer-generated block randomization sequence with concealed allocation. Study medications will be prepared by an independent anesthesiologist not involved in patient management or outcome assessment. Participants, clinical staff, outcome assessors, and statisticians will remain blinded to group allocation until completion of data analysis and database lock. The study drug and PCIA pump were uniformly prepared and labeled by the Department of Pharmacy according to the randomization numbers, and were identical in appearance and labeling. Both investigators and patients were blinded to group allocation. Unblinding was performed after completion and locking of the final dataset, at the time of the final statistical analysis.

### Data collection and record-keeping

The demographic and baseline characteristic data will be collected by screeners when the patients are recruited. Outcome assessors will measure clinical outcome measurement in 3 days after the operation. During the follow-up period, the participants’ incidence of complications will be investigated by a telephone call at 7 and 30 days after the operation.

Separate data collection forms for baseline information and postoperative follow-up were established using Feishu (Lark) multidimensional tables. Data were collected in groups, and an electronic database was subsequently created. Double data entry and verification were performed by two independent researchers. All personnel involved in anesthesia administration and data collection received standardized training. Intraoperative anesthesia depth was tightly controlled to ensure consistency between the two groups. Patients were properly instructed on wearing the wristbands to ensure data integrity.

### Sample size

Based on the primary endpoints of this study (total sleep time and deep sleep time postoperatively), the significance level was set at α = 0.05 (two-sided), and the statistical power (1-β) was set at 0.80, with reference to the preliminary experiment and relevant literature^8,12^. Assuming a mean difference of δ = 5% in total sleep efficiency (SE) between the two groups, with a standard deviation (SD) of 7%, the sample size was calculated using the formula for two independent samples t-test:

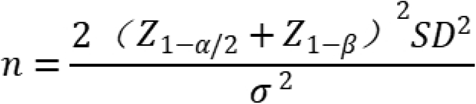

The calculation yielded n ≈ 31 patients per group.

Accounting for an estimated dropout rate of 10%, the planned total sample size for this study is 68 patients, with 34 patients in each group. This represents the minimum sample size required to achieve the intended statistical power.

### Statistical analysis

All analyses will follow the intention-to-treat principle. Continuous variables will be summarized as mean ± SD or median (IQR) as appropriate. Repeated sleep and pain measures will be analyzed using linear mixed-effects models with fixed effects for treatment group and time, and random effects for participants. Categorical variables will be compared using chi-square or Fisher’s exact tests. Missing data will be handled using multiple imputation. A two-sided p value < 0.05 will be considered statistically significant. Analyses will be performed using IBM SPSS 26 (version 26.0, IBM Corp., New York, NY, USA) software.

### Patient safety and public involvement

Any adverse events (defined as any functional lesion caused by the interventions, such as anesthetic allergy, poorly anesthetic effect during the operation, and so on) will be recorded. If any adverse event occurs, the doctor will provide the corresponding treatment to the patient, change, or even stop the anesthesia protocol. The adverse events will be immediately reported to the primary investigator and ethics committee to decide if the patient needs to withdraw from the trial. The trail conduct will be audited periodically in accordance with established standard operating procedures, and the auditing process will be conducted independently from both investigators and sponsors.

### Ethics and dissemination

The authors are accountable for all aspects of the work in ensuring that questions related to the accuracy or integrity of any part of the work are appropriately investigated and resolved. The study will be conducted in accordance with the Declaration of Helsinki and Good Clinical Practice guidelines. Ethical approval has been obtained from the Medical Ethical Committee of The Second People’s Hospital of Neijiang. Written informed consent will be obtained from all participants prior to enrollment. Study findings will be disseminated through peer-reviewed journal publications and academic conferences. Individual participant data will remain confidential and anonymized in all reports.

## Discussion

In this randomized, double-blind controlled trial of patients with breast carcinoma, we intend to compare the effects of Oxycodone and sufentanil on postoperative sleep quality and complications in patients after modified radical mastectomy. We will conduct this trial in strict accordance with the Consolidated Standards of Reporting Trials (CONSORT), and will be able to determine the effectiveness of general anesthesia. First of all, we estimate the optimal sample size to ensure adequate test performance. Furthermore, we ensure that this study is a genuinely randomized controlled trial through the full implementation of randomization and blindness. Sufentanil and oxycodone are both commonly used opioids for postoperative analgesia in the clinic. The anesthesia procedure employed a standardized, dosage-controlled combined intravenous-inhalation anesthesia protocol, with intraoperative monitoring of anesthesia depth to guide drug titration. In the evaluation index, a reasonable specification and a rigorous professional scale are designed as the assessment index of the primary outcomes, so as to avoid the subjective assessment and reduce the data distortion and deviation. Anesthesiologists with abundant clinical experience will perform the general anesthesia, The analgesic pump will be prepared by a designated resident physician, while the wearing of wristbands and data recording will be performed by an independent anesthesia nurse. Independence of personnel at each step was maintained to ensure blinding.

We hypothesized that, compared with sufentanil, oxycodone provides equivalent or superior postoperative analgesia while significantly improving postoperative sleep efficiency and reducing sleep fragmentation after modified radical mastectomy. Owing to its favourable adverse effect profile, oxycodone is increasingly used for perioperative analgesia. Previous studies have largely focused on its analgesic efficacy and side effects^3^, with limited evidence on its influence on sleep. While the unique advantages of oxycodone in visceral pain are well recognised^13^, we sought to investigate whether κ-opioid receptor activation improves postoperative sleep quality independently of its visceral analgesic effects and in the absence of visceral surgery. Polysomnography (PSG) remains the gold standard for sleep monitoring, but its use in postoperative patients is limited by poor compliance, patient discomfort, and interference with nursing care and mobilisation. In contrast, wearable devices enable unobtrusive, longitudinal monitoring under real-world conditions. At the group-comparison level, wearable-derived metrics such as total sleep time and sleep efficiency have demonstrated moderate-to-good agreement with PSG, particularly when used to assess relative changes rather than absolute sleep architecture^8,11^. This supports their application in randomized controlled trials focusing on between-group differences. We also aim to assess the concordance between subjective sleep experience and wearable device data.

By integrating a modern photoelectric device with a first-line analgesic, this interdisciplinary study addresses a gap in knowledge regarding κ-opioid receptor effects on postoperative sleep. We acknowledge two main limitations. First, this exploratory study used a relatively small sample size to conserve clinical resources. Second, PPG cannot fully replace the PSG, the established gold standard. Several methodological strategies were implemented to mitigate these limitations. Sleep parameters were analyzed within a predefined nocturnal window to reduce bias from prolonged postoperative bed rest. In addition, baseline-adjusted analyses were performed to account for inter-individual variability in habitual sleep patterns. Sensitivity analyses using alternative sleep window definitions yielded consistent results. Future studies with larger sample sizes are needed to investigate the effects of a broader range of perioperative drugs on sleep. Given the recognised importance of sleep, we hope these findings will inform perioperative sleep management, guide clinical analgesic selection, and improve patient recovery quality, support the growing role of wearable-derived digital biomarkers in perioperative research concurrently. While not a substitute for polysomnography, such tools provide a scalable and clinically practical approach to capturing sleep disruption in real-world surgical populations.

## Data Availability

For Study Protocols: No datasets were generated or analysed during the current study. All relevant data from this study will be made available upon study completion.

## Contributions

(I) Conception and design: Deng Qianyao, Liang Zheng; (II) Administrative support: Wu Ailing; (III) Provision of study materials or patients: Zheng Ju, Huang Lihua; (IV) Collection and assembly of data: Deng Qianyao, Hu Jie, Huang Lihua; (V) Data analysis and interpretation: Deng Qianyao, Zheng Ju, Liang Zheng; (VI) Manuscript writing: All authors; (VII) Final approval of manuscript: All authors.

